# Detecting radiographic sacroiliitis using deep learning with expert-level accuracy in axial spondyloarthritis

**DOI:** 10.1101/2020.05.19.20105304

**Authors:** Keno K. Bressem, Janis L. Vahldiek, Lisa C. Adams, Stefan M. Niehues, Hildrun Haibel, Valeria Rios Rodriguez, Murat Torgutalp, Mikhail Protopopov, Fabian Proft, Judith Rademacher, Joachim Sieper, Martin Rudwaleit, Bernd Hamm, Marcus R. Makowski, Kay Geert A. Hermann, Denis Poddubnyy

## Abstract

Conventional radiographs of sacroiliac joints from two independent cohorts of patients with axial spondyloarthritis (axSpA) were used to develop and validate an artificial neural network for the detection of definite radiographic sacroiliitis as a manifestation of the disease. The first cohort consisting of 1669 radiographs labelled by three experts was used for training. From the second cohort 100 radiographs labelled by two experts were randomly selected for the test dataset. The neural network achieved an excellent performance in detection of definite radiographic sacroiliitis with an area under the receiver operating characteristics curve of 0.97 and 0.96 for the validation and test datasets, respectively. Sensitivity and specificity for the cut-off weighting both measurements equally were 0.90 and 0.93 for the validation and 0.87 and 0.97 for the test set. The Cohen’s kappa between the neural network and the reference judgements were 0.80 for both validation and test sets, and the absolute agreement on the classification yielded 91% and 90%, respectively.

## INTRODUCTION

Axial spondyloarthritis (axSpA) is a chronic inflammatory disorder primarily affecting axial skeleton – sacroiliac joints and spine. Detection of radiographic sacroiliitis has been for many years the only possibility to establish a definite diagnosis of the disease prior to development of structural damage in the spine. The presence of definite radiographic sacroiliitis (defined as sacroiliitis of at least grade 2 bilaterally or at least grade 3 unilaterally) is also an obligatory criterion in the modified New York criteria for ankylosing spondylitis (AS).^1^ Although magnetic resonance imaging (MRI) of sacroiliac joints nowadays allows to diagnose axSpA earlier, definite radiographic sacroiliitis can be detected at the time-point of the diagnosis already in about 33% of the patients with symptom duration of up to one year and about 50% of the patients with symptom duration of 2 to 3 years.^2^ Conventional radiography of the sacroiliac joints is still recommended as the first imaging method if axSpA is suspected.^3^ Furthermore, radiographic sacroiliitis is included – together with sacroiliitis on MRI – in the Assessment of Spondyloarthritis International Society (ASAS) classification criteria for axSpA.^4^ Depending on the presence or absence of definite radiographic sacroiliitis, axSpA can be classified either as radiographic axSpA (r-axSpA, synonymous to AS) or non-radiographic axSpA (nr-axSpA).^5^ Such a classification might be relevant for both clinical practice (currently, labels for biological disease modifying antirheumatic drugs – bDMARDs are different for AS and nr-axSpA) and for research (i.e., stratification or selection of patients in a clinical trial).

Although conventional radiography of sacroiliac joints still plays an important role in clinical practice and for clinical trials, the reliability of radiographic sacroiliitis assessment has been reported in a number of studies as mostly poor, even if performed by expert readers.^6-10^ Furthermore, it was shown that untrained local readers perform worse than expert readers specialised in SpA.^10^ One possible solution to achieve a comparable high accuracy as an expert on detection of radiographic sacroiliitis, even in non-specialised clinics, could be development of an artificial intelligence-based model for analysis of radiographs. Artificial intelligence-based or deep learning models have already accomplished remarkable results in the classification of medical and non-medical data. For example, neural networks have been trained to detect breast cancer in mammographs, to classify skin cancer or to label chest radiographs.^11-13^ Common to all these studies was that they did not develop a de novo model, but rather applied a transfer learning approach using a pre-trained network. This allows knowledge of pre-trained models from non-medical fields to be used for a new visual task, effectively reducing the amount of data required for training while increasing the accuracy of the models.

In the present study, we therefore aimed to develop and to validate a deep learning artificial neural network for the detection of definite radiographic sacroiliitis using centrally scored images from two observational cohort studies.

## METHODS

### Cohort description

In this project, we used imaging data from two independent sources: 1) <u>P</u>atients With Axial Spondyloarthritis: Multic<u>o</u>untry Registry <u>of</u> Clinical Characteristics (PROOF), and 2) German Spondyloarthritis Inception Cohort (GESPIC).

PROOF is an ongoing study conducted in clinical practices in 29 countries that includes 2170 adult patients diagnosed with axSpA (non-radiographic or radiographic) ≤12 months before study enrolment and fulfilling the ASAS classification criteria for axSpA. In 1669 patients, radiographs of sacroiliac joints were available for central reading.

GESPIC is a multicentre inception cohort study conducted in Germany that includes 646 patients with SpA.^14^ In 525 patients, radiographs of sacroiliac joints were available for central reading.

### Assessment of radiographic sacroiliitis

Baseline radiographs of sacroiliac joints were collected, digitized if necessary, anonymized and subsequently centrally scored by trained and calibrated readers according to the grading system of the modified New York criteria:^1^

Grade 0 normal;

Grade 1 suspicious changes;

Grade 2 minimal abnormality: small localized areas with erosion or sclerosis, without alteration in the joint width;

Grade 3 unequivocal abnormality: moderate or advanced sacroiliitis with erosions, evidence of sclerosis, widening, narrowing, or partial ankylosis;

Grade 4 severe abnormality: total ankylosis.

In the PROOF study, images were first assessed by the local readers, then by the central reader 1 (DP) who was blinded for the local assessment. In case of a disagreement on the presence of definite radiographic sacroiliitis (grade ≥2 bilaterally or grade ≥3 unilaterally) between the local and central reader 1, the radiograph was evaluated by central reader 2 (HH) who was blinded to the previous assessments. The final decision on the presence of definite radiographic sacroiliitis and, therefore, on classification as nr-axSpA or r-axSpA was made based on the decision of two of the three readers.

In GESPIC, no local reading of radiographs was demanded; all collected images were scored independently by two trained and calibrated central readers (VRR and MT).

### Image selection and pre-processing

The PROOF dataset consisted of 1669 radiographs of sacroiliac joints in DICOM (Digital Imaging and Communications in Medicine) format, varying in size, resolution and quality. 18 images were excluded due to very poor image quality (Figure 1). The Horos Project DICOM Viewer (version 4.0.0, www.horosproject.org) was used to manually adjust the greyscale-levels of all images and to convert them afterwards to the Tagged Image File Format (TIFF). If other body parts such as the thoracic spine were included in the image, it was manually cropped to the pelvis. The final data set for building the model consisted out of 1651 individual patient images and was split randomly into a training (85% - 1404 radiographs) and validation data set (15% - 247 radiographs).

**Figure 1.**
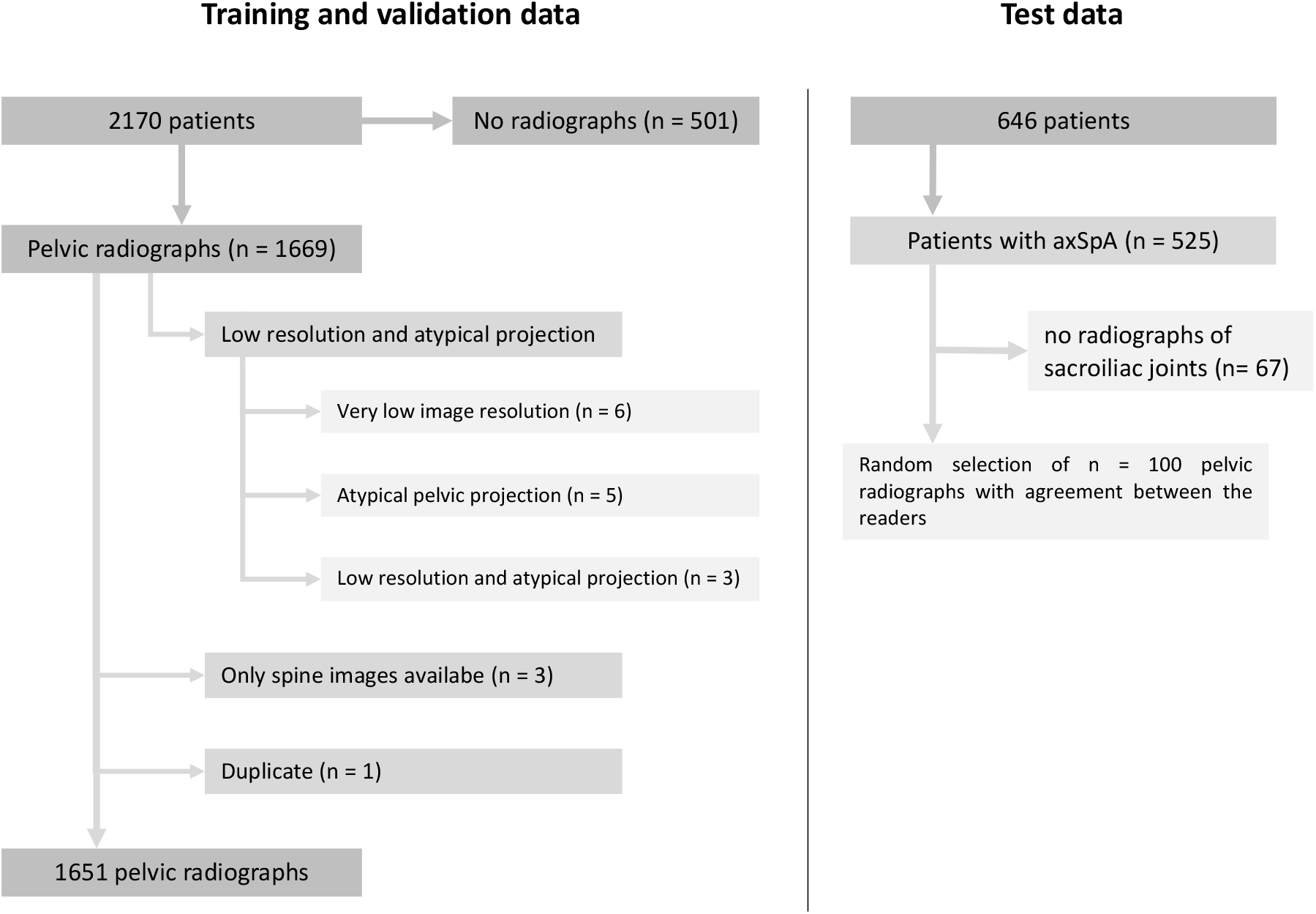
shows a flowchart for selection of cases from the PROOF (training and validation set) and GESPIC (test set) studies. Overall, 18 radiographs had to be excluded from the PROOF study, mostly because of low image resolution or atypical pelvic projections of the radiographs, which were not in accordance to standardised pelvic radiographs. For the test data set, 100 images with the agreement on the presence or absence of definite radiographic sacroiliitis were randomly selected and subsequently used for the final model evaluation.

For testing the generalisability across data sets, 100 radiographs of sacroiliac joints with agreement of both readers on the presence or absence of definite radiographic sacroiliitis were randomly selected from the 525 baseline radiographs of GESPIC and then pre-processed exactly as the training and validation data sets.

### Model Training

Model training was performed on a dedicated Ubuntu 18.04 Workstation with two Nvidia GeForce RTX 2080ti Graphic cards as well as on a GPU node of the Berlin institute of health (BIH) high performance computing cluster using four Nvidia Tesla V100 Graphic Cards. All model training was mainly performed using Python (version 3.7), with the fastAI application programming interface which is built on top of PyTorch.^15,16^

As base model, a convolutional neural network (ResNet-50 architecture), pre-trained on the image net data set, consisting of over 14 million images, was used.^17^ The images were augmented prior to training through various transformations consisting of flipping, rotation of up to 10°, magnification of up to 1.1, lighting variations and warping. We further utilised the mix-up method during training, originally introduced by Zhang et al,^18^ in which images of different classes (nr-axSpA and r-axSpA) are combined during training to reduce memorisation of noisy labels and increase overall model robustness. As a loss function, we utilised cross entropy label smoothing, which reduced high-confidence predictions of the models, thus supporting regularisation and avoiding overfitting with subsequent better generalisation of the models on new data (e.g. test-data set). Model training was performed with discriminative learning rates and a progressive resizing approach, starting with images-sizes of 224×224 pixels (which is the default input-size for the ImageNet pretrained ResNet-50) and then increasing the resolution to 512×512 pixels and 768×768 pixels.^19^ During training, first only the last two classification layers of the model were trained, with the weights of the other network-layers remaining frozen. A total of 100 epochs were trained, monitoring the area under the receiver-operating characteristics curve (AUROC) on the validation data set, saving the models weights on every improvement. After 100 epochs, the weights of the model with the highest AUROC value were re-loaded, the model was unfrozen and again trained for 100 epochs, while monitoring the AUROC and saving the weights at every improvement. This approach was repeated for all image resolutions. The size of the mini batches was 64 for 224×224 pixels, 32 for 512×512 pixels and 84 for 768×768 pixels. The training for lower resolutions could be performed at our local Workstation, while for 768×768 pixels the BIH high performance cluster was used. Overall, model training took approximately 24 hours on our local machine and about an additional 6 hours on the cluster. After training, activation maps were created to verify that the model used the sacroiliac joints to determine whether definite radiographic sacroiliitis was present.

### Statistical analysis

Statistical analysis was performed using the R statistical environment (version 3.6), the “tidyverse”, “ROCR” and “irr” libraries.^20-23^ Raw predictions of the model on the validation data set as well as on the test data set using an image resolution of 768×768 pixels were exported from the python environment as comma separated values and imported into “R”. ROC-curves and precision recall curves were plotted and the area under the curve was calculated. A ROC analysis was then performed to identify different prediction cut-offs. Sensitivity and specificity of the cut-offs favouring sensitivity, favouring specificity or balancing both were calculated. Confusion matrices were constructed using the prior defined cut-offs. Cohen’s kappa and the percentage absolute agreement was used to assess the agreement between human and machine. 95% confidence intervals for calculated kappa values were estimated using bootstrapping with 1000 repetitions. A p-value of < 0.05 was considered statistically significant.

### Patient and Public Involvement

This study as well as both parental cohorts were done without any formal patient/patient organization involvement in study design, development of patient relevant outcomes, interpretation of results, or the writing or editing of the manuscript.

### Ethics approval

Both PROOF and GESPIC cohorts were approved by the local ethics committees of each study centre in accordance with the local laws and regulations and is being conducted in accordance with the Declaration of Helsinki and Good Clinical Practice. GESPIC was additionally approved by a central ethics committee of the coordinating centre. Written informed consent was obtained from all patients.

## RESULTS

Definite radiographic sacroiliitis was present in 911 (64.9%) patients of the training set (PROOF, n=1404), in 160 (64.8%) patients of the validation set (PROOF, n=247) and in 54 (54%) patients of the independent test set (GESPIC, n=100). In a total of 392 (27.9%) and 67 (27.1%) patients in the training and validation sets, there was a discrepancy between the local and the central reader 1 that automatically resulted in the involvement of the central reader 2. A total of 160 (11.4%) and 39 (15.8%) patients in the training and validation sets were re-classified after the central reading, meaning the rating from both central readers differed from the rating of the local reader.

### Model performance in the validation data set

On the validation data set, the model achieved an excellent performance. The ROC analysis showed an area under the curve (AUROC) of 0.971. For the precision-recall curve a value of 0.989 could be calculated. Both the local and central expert readers remained below the ROC and PR curves and were therefore outperformed by the accuracy of the model. We propose three cut-offs to convert the floating-point predictions into integer values, where a 1 represents the presence of definite radiographic sacroiliitis and 0 the absence. Cut-offs weighted sensitivity over specificity and specificity over sensitivity, aiming to find the optimal balance between both measurements (defined as maximal sum between sensitivity and specificity). The first cut-off value, which favours sensitivity over specificity, was calculated to be 0.606, resulting in a sensitivity of 0.994 and a specificity of 0.667 for the detection of radiographic SpA. The second cut-off, which favoured specificity over sensitivity, was 0.814, resulting in a sensitivity of 0.656 and a specificity of 1. The third cut-off was 0.706, resulting in a sensitivity of 0.9 and a specificity of 0.931. The human local reader had a sensitivity of 0.844 and a specificity of 0.839. The expert central reader had a higher sensitivity than the local reader with 0.975, but at the cost of a lower specificity of 0.724. The performance of the model is also shown in Figure 2 in the form of ROC curves and precision recall curves and Table 1 as confusion matrices with kappa-values and values of absolute agreement.

**Table 1.**
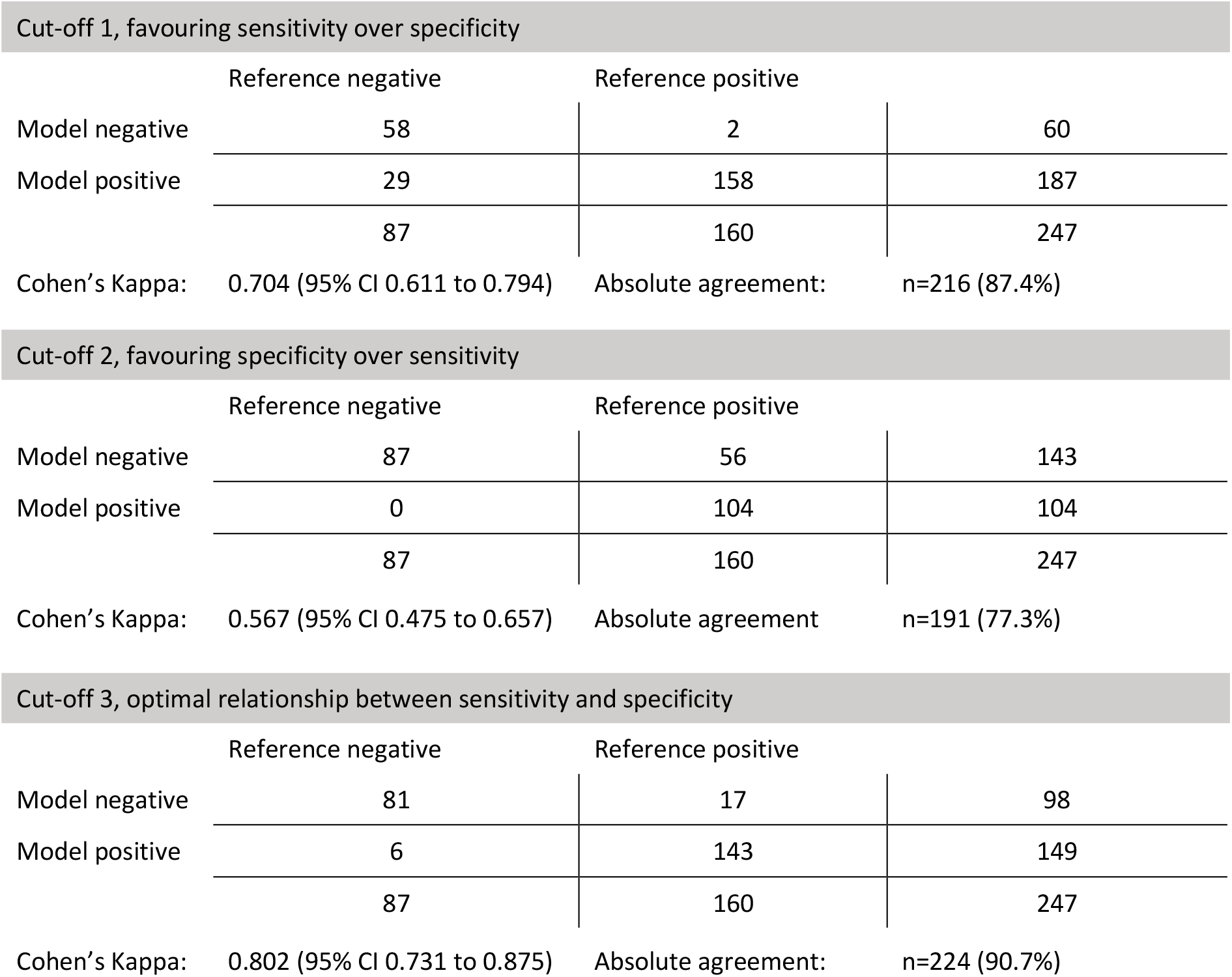
provides confusion matrices for the three proposed cut-offs for the model-predictions for the presence of definite radiographic sacroiliitis on the validation data set alongside the agreement between ratings measured by Cohen’s Kappa and the absolute percentage agreement. Cut-off 1 weights sensitivity more than specificity, while cut-off 2 weights specificity more than sensitivity and cut-off 3 aims to be the optimal trade-off between sensitivity and specificity.

**Figure 2.**
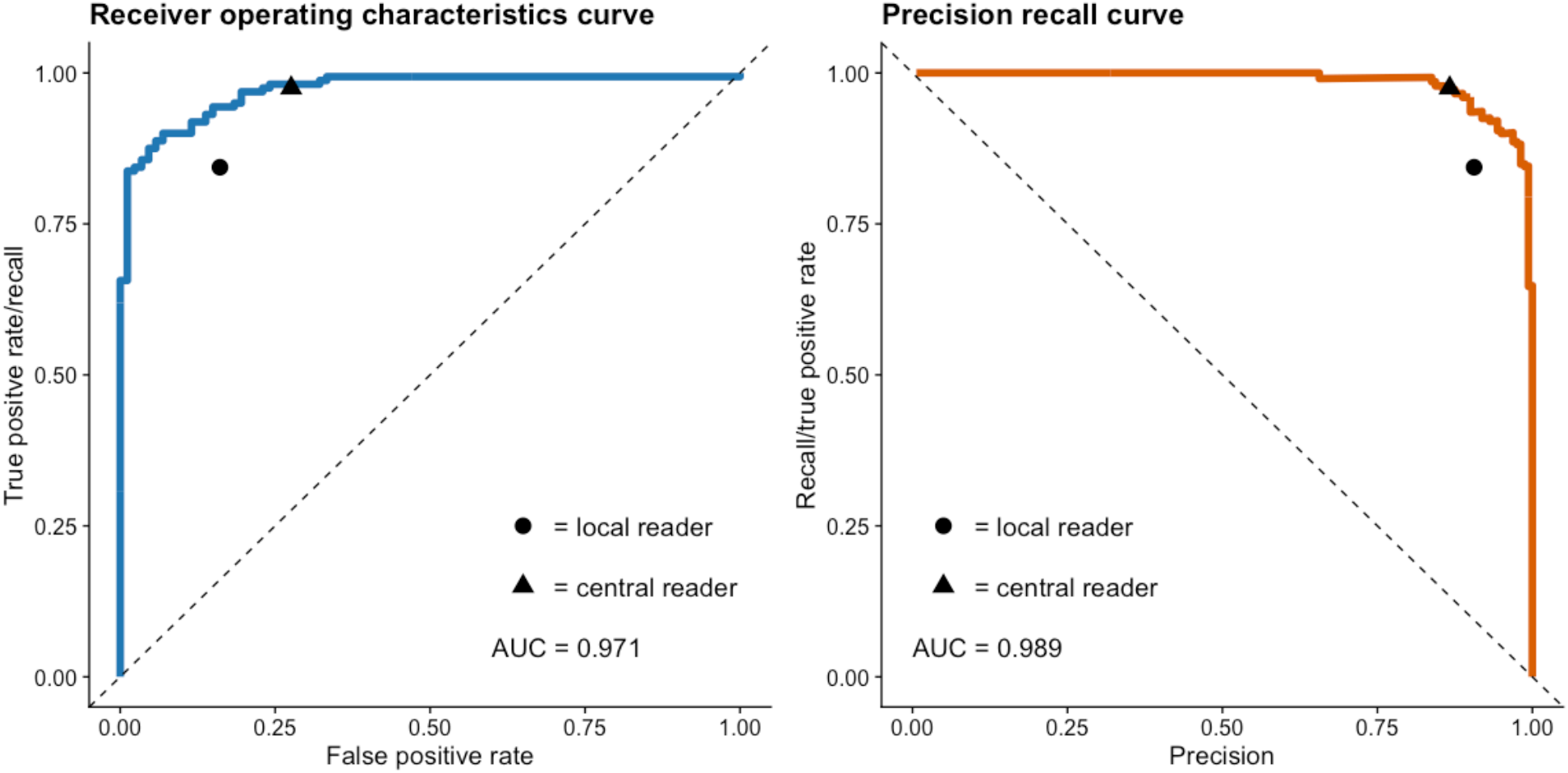
shows the receiver operation characteristics curve and the precision recall curve for the model performance in detection of definite radiographic sacroiliitis (classification as non-radiographic or radiographic axSpA) on the validation data set as well as the corresponding area under the curve (AUC). Individual values for both expert readers are displayed as a triangle or period.

The reference was the presence of definite radiographic sacroiliitis according to the majority decision of three readers in the PROOF cohort.

With the cut-off with balanced sensitivity and specificity, the neural network achieved a kappa of 0.80 (95% CI 0.73 to 0.88) and an absolute agreement of 91% on the final classification. This was superior to the agreement between the local reader judgement and the final classification (Cohen’s kappa = 0.66, 95%CI 0.57 to 0.76; absolute agreement 84%) and even better than the agreement between the central reader judgement and the final classification (Cohen’s kappa = 0.74, 95%CI 0.65 to 0.83 absolute agreement 89%).

### Model performance in the independent data set

In the test data set, the model performance was slightly worse than on the validation data set with an AUROC-value of 0.961 and an area under the precision recall curve (AUPRC) value of 0.979. Again, we propose three cut-offs: The first cut-off, which weighs sensitivity over specificity, was 0.58, yielding a sensitivity of 1 and a specificity of 0.478. The second cut-off, which weighs specificity over sensitivity, was 0.834, based on which a specificity of 1 and a sensitivity of 0.574 could be achieved. The optimal performance for both performance measurements could be achieved with a cut-off of 0.791, resulting in a sensitivity of 0.87 and specificity of 0.966. Figure 3 shows the ROC- and precision-recall curves for the model performance on the test-data set. Figure 4 demonstrates the different values for sensitivity and specificity defined for different cut-offs on the test and validation data set. Table 2 provides confusion matrices for the three proposed cut-offs on the test data set alongside evaluation of agreement in the form of Cohen’s kappa and percentage agreement. Importantly, in this independent test set, almost the same performance of the model with balanced sensitivity and specificity could be achieved as in the validation data set: Cohen’s kappa = 0.80, absolute agreement of 90%.

**Table 2.** provides confusion matrices for the three proposed cut-offs for the raw model-predictions for the presence of definite radiographic sacroiliitis on the test data set alongside the agreement between ratings measured by Cohens Kappa and the absolute percentage agreement. Cut-off 1 weights sensitivity more than specificity, while cut-off 2 weights specificity more than sensitivity and cut-off 3 aims to be the optimal trade-off between sensitivity and specificity. The reference was the agreement of two central readers on the presence of definite radiographic sacroiliitis in the GESPIC cohort.

| Cut-off 1, favouring sensitivity over specificity |  |  |  |
| --- | --- | --- | --- |
|  | Reference negative | Reference positive |  |
| Model negative | 24 | 1 | 25 |
| Model positive | 22 | 53 | 75 |
|  | 46 | 54 | 100 |
| Cohens Kappa: | 0.521 (95% CI 0.372-0.672) | Absolute agreement | n= 77 (77%) |

| Cut-off 2, favouring specificity over sensitivity |  |  |  |
| --- | --- | --- | --- |
|  | Reference negative | Reference positive |  |
| Model negative | 46 | 24 | 70 |
| Model positive | 0 | 30 | 30 |
|  | 46 | 54 | 100 |
| Cohens Kappa: | 0.535 (95% CI 0.389-0.680) | Absolute agreement: | n=76 (76%) |

| Cut-off 3, optimal relationship between sensitivity and specificity |  |  |  |
| --- | --- | --- | --- |
|  | Reference negative | Reference positive |  |
| Model negative | 44 | 8 | 52 |
| Model positive | 2 | 46 | 48 |
|  | 46 | 54 | 100 |
| Cohens Kappa: | 0.801 (95% CI 0.688-0.919) | Absolute agreement: | n=90 (90%) |

**Figure 3.**
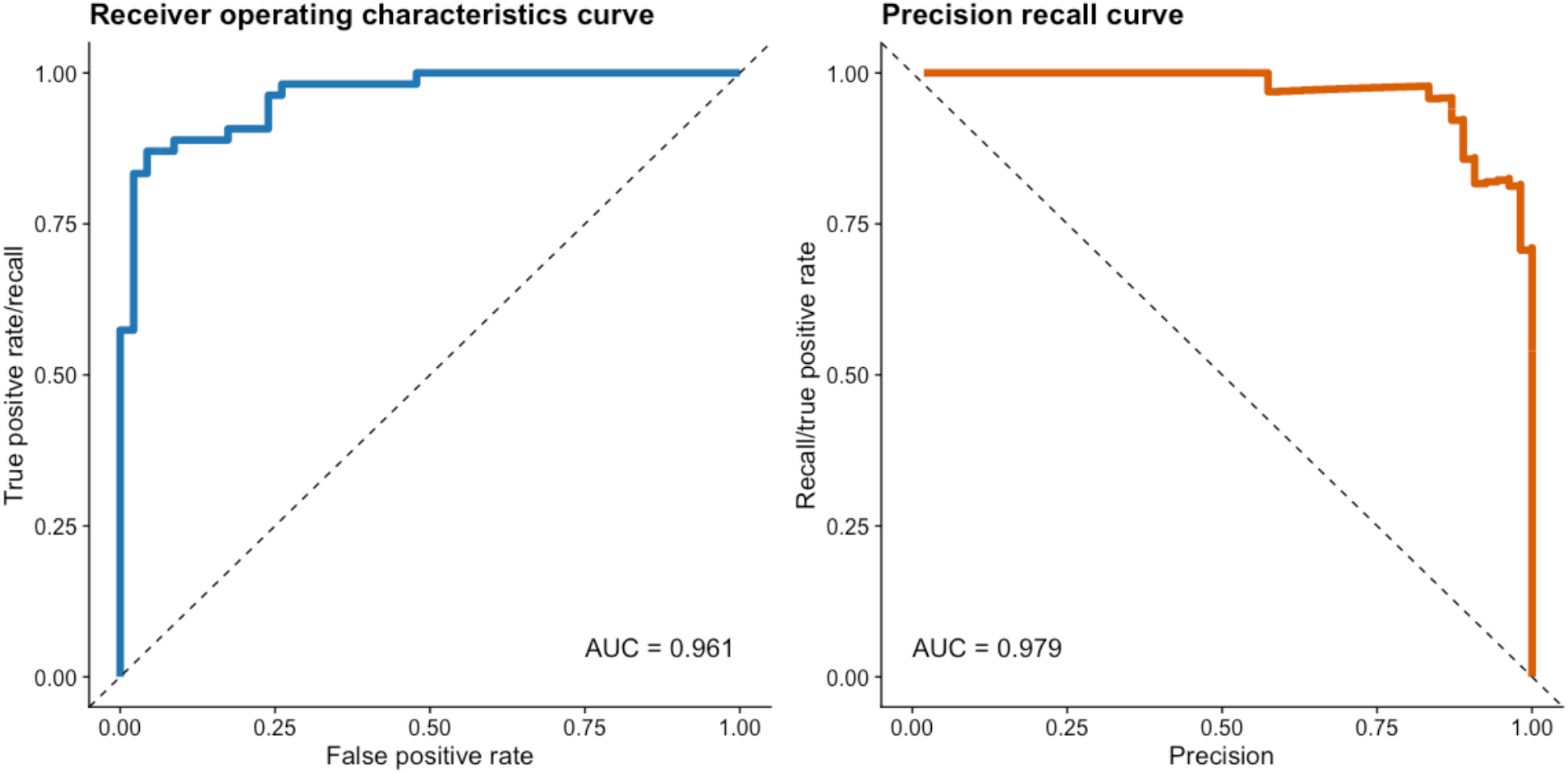
shows the receiver operation characteristics curve and the precision recall curve for the model performance in detection of definite radiographic sacroiliitis (classification as non-radiographic or radiographic axSpA) on the test data set alongside respective area under the curve (AUC). Since the reference standard here was the agreement of two independent readers, a presentation of the accuracy of these readers was not performed.

**Figure 4.**
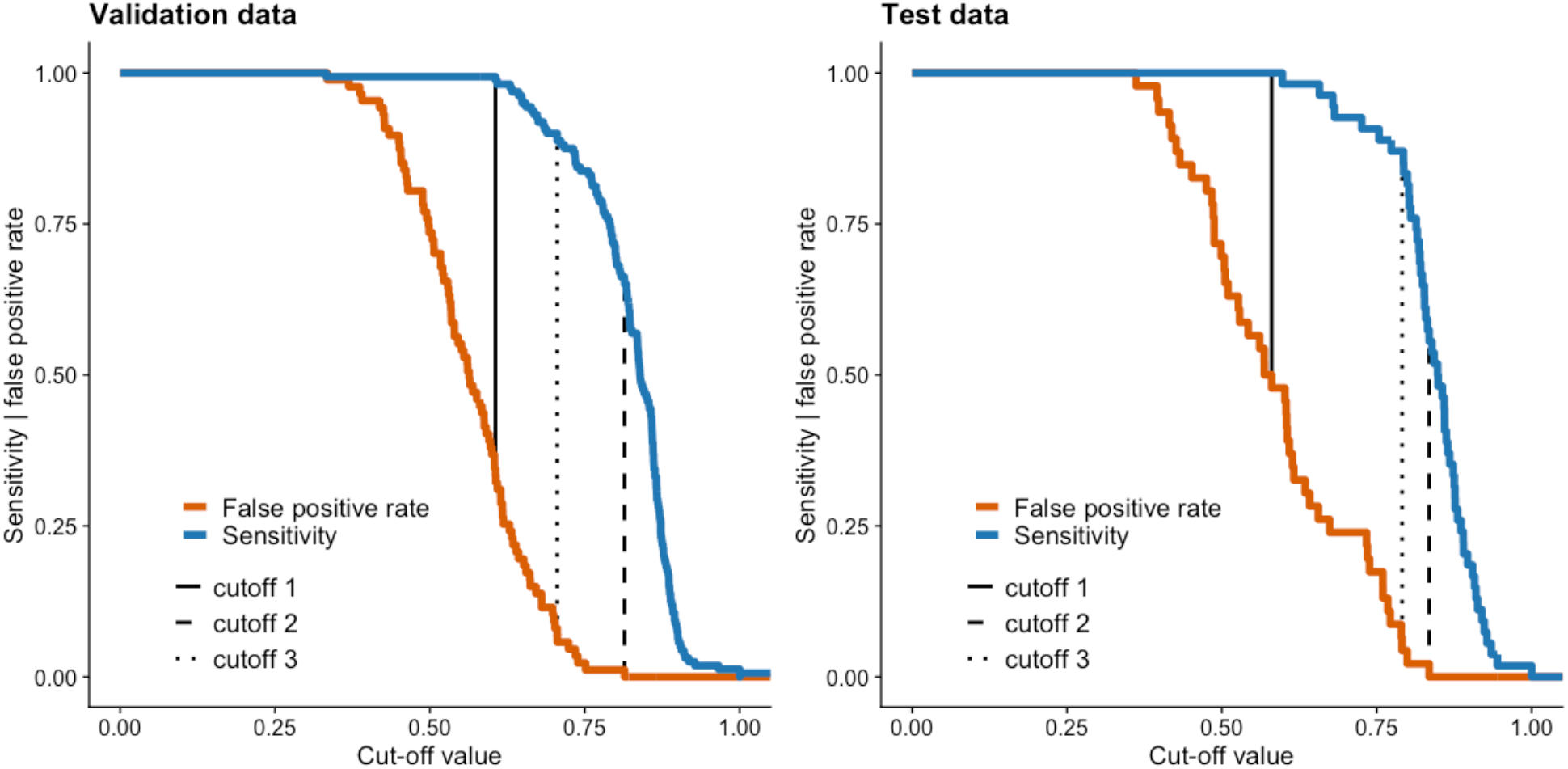
shows the sensitivity and 1-specificity (false positive rate) on the test and validation data set with varying cut-off values for the model predictions on the presence of radiographic sacroiliitis (classification as non-radiographic or radiographic axSpA). We analysed three cut-off values, indicated by vertical dashed lines. Cut-off 1 weights sensitivity over specificity, cut-off 2 weights specificity over sensitivity and cut-off 3 aims to be the optimal balance between the two performance measurements.

Figure 5 shows activation maps of the neural network for correct and incorrect predictions on the test-and validation data sets.

**Figure 5.**
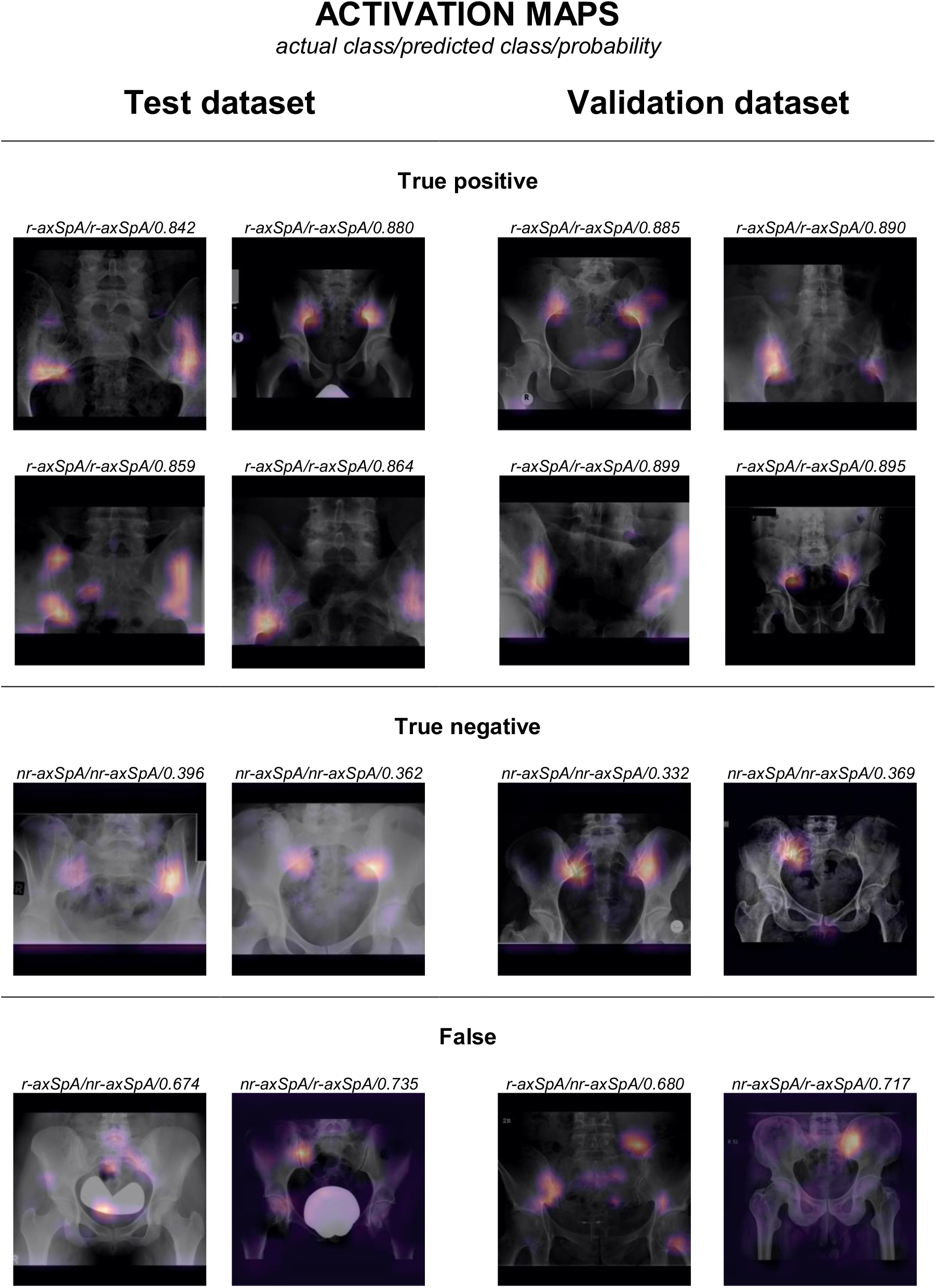
shows activation maps of the model on example images taken from the validation and test data sets. It can be seen, that the sacroiliac joints are the most decisive for the model’s predictions, but the lumbo-sacral joint and the hip joints also appear to be partially relevant for some predictions.

## DISCUSSION

In this study, we successfully developed and tested an artificial intelligence model for the detection of radiographic sacroiliitis on conventional radiographs. With the developed model, we could even achieve a super-human accuracy on the validation data. Furthermore, we were able to demonstrate the generalisability of our model on a test data set of novel data, achieving a performance at least comparable to that of two human experts.

Although in industrialized countries magnetic resonance imaging is increasingly used for the detection of sacroiliitis, radiographs are still of importance. Especially in developing countries radiographs are the main imaging procedure due to a low availability and high associated costs of magnetic resonance imaging. Detection of definite radiographic sacroiliitis is important for both diagnosis and classification of axSpA. At the same time, a poor reliability of sacroiliitis detection by conventional radiographs is well known.^6-10^ In the present study we used a large and unique dataset to train, to validate, and to test the model. The resulting performance was at least as good as (but most likely better than) the performance of an experienced reader with expertise in radiographic sacroiliitis assessment. The neural network could achieve almost the same level of performance in both validation and training sets indicating high level of reliability and robustness of the model. The developed model can be applied, therefore, as an additional diagnostic aid in clinical practice and as a classification tool in research projects involving patients with axSpA.

Neural networks have already been applied to a variety of medical imaging data, including radiographs but, to our knowledge, not for the detection of spondyloarthritis.^11-13,24^ However, a low generalisability, i.e. poor performance of the models on new data, is an important challenge in training neural networks. A new meta-analysis on ‘deep learning performance against healthcare professionals’ by *Kim et al*. revealed methodological shortcomings that are present in many published studies on deep learning in medicine.^25^ They criticized that many studies either did not compare the performance of their model to that of a human domain expert, or evaluated the performance of their model on a dataset different from the dataset used to evaluate human performance, so that they achieve incorrectly high accuracies, mainly due to over-adaptation, and consequently have a low generalisability.^25^ Similar observations were made by *Yao et al*., who showed that although 155 studies on deep learning in medicine have been published, they often lack external validation data.^26^ However, the use of external validation data is an important measure to proof generalisability. It has been shown that medical computer vision models adapt poorly to the use of different scanners or imaging protocols, and the lack of external validation is likely to result in poor generalisability of the model to new data.^27^ In a recent study, *McKinney et al* evaluated the performance of a neural network for the detection of breast cancer in mammographs, surpassing human performance.^11^ They used different data sets from different studies to train and test their developed models and were thus able to demonstrate sufficient generalisability of their developed models.

Similar to their approach, we also used a heterogeneous training data set with radiographs from different imaging sites and achieved a good generalisability of the developed model, with the performance being only slightly inferior on the test data, which was independent in terms of patients and readers, as compared to the validation data.^18,28,29^ While the heterogeneity of our training data set already reduced the risk of overfitting on systematic image noise, e.g. to device specific image features, we further increased generalisability by applying progressive resizing and the integration of mix-up as well label-smoothing into model-training.

Our study has some limitations. First, the reference for the training of the model was the judgement of a limited number (2 or, in the case of discrepancy, 2 out of 3) of human readers. Although both central readers in the PROOF study had many years of experience in reading of radiographs of sacroiliac joints, the complex anatomy of sacroiliac joints and heterogeneity of radiographic techniques and quality have brought an inevitable portion of uncertainty in the final classification used as a reference. In the independent dataset, we selected only cases where both readers agreed to be the reference standard for the evaluation of the model. This approach was chosen because we believe that these cases are most likely true positive or true negative ones. Other approaches, such as evaluating all cases, in which the readers disagreed, to be negative, were discarded as we saw a risk of biasing the test data set and thereby distorting the evaluation of the model performance. Remarkably, despite all the uncertainty related to the assessment of radiographic sacroiliitis, a high level of agreement between the neural network’s judgement and the consensus judgement by human could be achieved in both validation and test sets.

To further evaluate the model performance, we believe a prospective study is needed since the data we used is from two studies not primary designed for training of a deep neural network. Thus, a selection bias might be present in our data.

## Conclusion

Convolutional neural networks can reliably detect definite radiographic sacroiliitis with a performance similar or even superior to that of a human expert reader.

## Data Availability

Data are available upon reasonable request.

## ACKNOWLEDGMENTS

LCA is grateful for her participation in the BIH Charité–Junior Clinician and Clinician Scientist Program and KKB is grateful for his participation in the BIH Charité Digital Clinician Scientist Program all funded by the Charité–Universitätsmedizin Berlin and the Berlin Institute of Health. We would like to thank the Berlin Institute of Health (BIH) for providing access to their high-performance computing cluster for our analysis.

## AUTHOR CONTRIBUTIONS

Conceived and designed the study: KKB, JLV and DP; Data processing and extraction: KKB, JLV and DP, HH, JS, MR, KGAH; Analyzed the data: KKB, JLV, MR, JS, HH and DP; Training of deep neural networks: KKB, JLV; Drafting of the manuscript: KKB, JLV, LCA, DP; Commented on the manuscript, suggested/made revisions of important intellectual content and approved the final version: KKB, JLV, LCA, SMN, HH, VRR, MT, MP, FP, JR, JS, MR, BH, MRM, KGAH, DP; Administrative and technical support: DP, SMN, BH and MRM.

